# Preoperative CT-Based Habitat Radiomics Classifiers Predict Recurrence in Non-Small Cell Lung Cancer

**DOI:** 10.64898/2026.04.14.26350899

**Authors:** Oya Altinok, Wai Lone J. Ho, Lary Robinson, Dmitry Goldgof, Lawrence O. Hall, Albert Guvenis, Matthew B. Schabath

## Abstract

**Objectives:** Among surgically resected non-small cell lung cancer (NSCLC) patients with similar stage and histopathological characteristics, there is variability in patient outcomes which highlights urgency of identifying biomarkers to predict recurrence. The goal of this study was to systematically develop a pre-surgical CT-based habitat-based radiomics classifier to predict recurrence-of-risk in NSCLC.

**Methods:** This study included 293 NSCLC patients with surgically resected stage IA–IIIA disease that were randomly divided into a training (n = 195) and test cohorts (n = 98). From pre-surgical CT images, tumor habitats were generated using two-level unsupervised clustering and then radiomic features were calculated from the intratumoral region and habitat-defined subregions. Using ridge-regularized logistic regression, separate classifiers were developed to predict 3-year recurrence using intratumoral radiomics, habitat-based radiomics, and a combined model (intratumoral and habitat) which was generated using a stacked learning framework. For each classifier, probability of recurrence was calculated for each patient then numerous statistical and machine learning approaches were utilized to stratify patients for recurrence-free survival.

**Results:** The combined radiomics classifier yielded a superior AUC (0.82) compared to the intratumoral (AUC = 0.75) and habitat radiomics (AUC = 0.81) models. When the classifiers were used to stratify high- versus low-risk patients utilizing a cut-point identified by decision tree analysis, high-risk patients were yielded the largest risk estimate (HR = 8.43; 95% CI 2.47 – 28.81) compared to the habitat (HR = 5.41; 95% CI 2.08 – 14.09) and intratumoral radiomics (HR = 3.54; 95% CI 1.45 – 8.66) models. SHAP analyses indicated that habitat-derived information contributed most strongly to recurrence prediction.

**Conclusions:** This study revealed that habitat-based radiomics provided superior statistical performance than intratumoral radiomics for predicting recurrence in NSCLC.

## Introduction

Lung cancer is the leading cause of cancer-related mortality in the United States [1]. Among patients diagnosed with surgically resectable early-stage non-small cell lung cancer (NSCLC), 3-year recurrence occurs in approximately 13% of patients with stage I disease and 42% for stage II [2–4]. However, patients with similar stage and histopathological characteristics can experience markedly different survival outcomes [5]. This variability in patient outcomes highlights urgency of developing precise recurrence risk stratification approaches and to generate precise biomarkers to identify early-stage NSCLC patients at high risk-of-recurrence.

Analysis of standard-of-care medical imaging (e.g., computed tomography [CT]) utilizing radiomic features has emerged as a powerful approach to identify image-based biomarkers across the cancer control continuum including predicting survival outcomes including risk-of-recurrence [6–11]. However, prior studies that leverage radiomics to predict survival outcomes [9–11] only utilized intratumoral radiomics which captures features from the tumor as a singular homogeneous region-of-interest. While intratumoral measurements have predictive utility, this approach has limitations because it does not account for the intratumoral spatial heterogeneity, which has a critical role in tumor aggressiveness and disease recurrence [12–14]. As such, habitat-based analyses can be utilized to explicitly identify intratumoral spatial heterogeneity by partitioning tumors into distinct subregions and then calculating radiomic features from each subregion [15,16]. As radiomics capture the pathophysiological underpinnings of a region-of-region [17], habitat measurements are capturing biological heterogeneity and microenvironment complexity [18]. In lung cancer, habitat radiomics have been shown to predict molecular alterations [19], tumor invasiveness patterns [20], lymph node metastasis [21], and distant metastasis [22]. A recent study utilized a habitat imaging framework by integrating CT and ^18^F-FDG PET imaging to predict recurrence in NSCLC [23]. However, the added complexity and limited availability of multimodal imaging highlight the need for habitat-based approaches that can be implemented using routinely acquired CT alone. Collectively, these studies support the biological and clinical relevance of habitat imaging features.

The goal of this study was to systematically evaluate pre-treatment CT-based habitat-based radiomics and develop a classifier to predict recurrence-of-risk in surgically resected NSCLC. As part of this systematic evaluation framework, multiple habitat feature aggregation strategies were compared which collapse habitat-level feature vectors into the best statistically performing patient-level representation. Next, classifiers to predict recurrence were developed based on using only intratumoral radiomics, only habitat-based radiomics, and a combined model which concatenated the intratumoral and habitat radiomic models. For each classifier, probability of recurrence was calculated for each patient then numerous statistical approaches (e.g., median, tertile) and machine learning methods were utilized to identify the most informative thresholds to stratify patients into high- and low-risk of recurrence. Overall, this framework that utilizes standard-of-care pre-treatment medical images and data could have clinical utility to enable postoperative surveillance and adjuvant treatment clinical decision-making.

## Materials and Methods

### Study Population and Patient Data

Details of this study have been published elsewhere [24] and the overall workflow of this study is illustrated in Figure 1. In brief, pre-surgical clinical data and contrast-enhanced CT images were derived from a cohort of 708 patients diagnosed with primary NSCLC at the H. Lee Moffitt Cancer Center & Research Institute. Patients were included if they had stage IA–IIIA disease and underwent surgical resection as first-line treatment. The pre-surgical CT images were obtained up to 6 months before surgery. Patients were excluded from the analysis if they did not have a minimum of follow-up of 3 years, received preoperative chemotherapy or radiotherapy, or had multiple lung nodules. The final analytical sample size was 293 patients which was randomly divided into a training set (n = 195) and test set (n = 98) using stratified random sampling to maintain comparable recurrence distributions. The training cohort was used for feature selection and model training and performance evaluations were conducted in the unseen test cohort.

**Figure 1.**
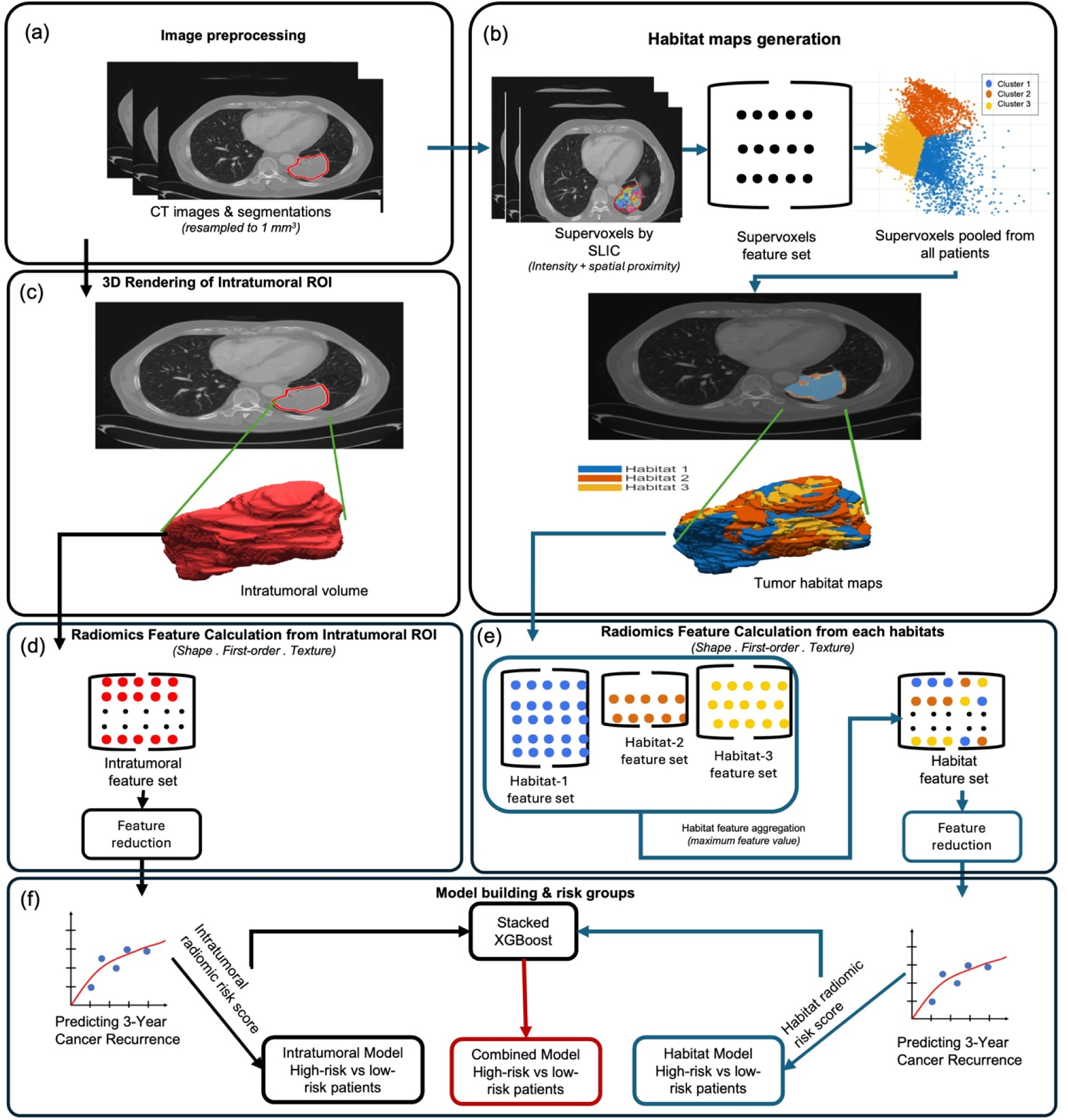
Workflow of the systematic evaluation framework deployed in this study. Pre-treatment contrast-enhanced CT images were used to delineate tumor regions of interest (ROI) and resampled to an isotropic resolution of 1 mm³ (a). Within each resampled tumor ROI, supervoxels were generated using the Simple Linear Iterative Clustering (SLIC) algorithm based on voxel-level intensity and spatial proximity. First-order radiomic features were calculated from each supervoxel and pooled across all patients for population-level K-means clustering, resulting in distinct tumor habitats. The resulting habitat labels were mapped back to individual tumors to generate three-dimensional patient-specific habitat maps (b). The resampled intratumoral ROI and corresponding tumor mask are shown as three-dimensional volumes for visualization (c). Radiomic features capturing shape, first-order statistics, and texture were calculated from the intratumoral ROI (d) and separately from each identified habitat (e). Habitat-level features were aggregated using a maximum-value strategy to obtain a fixed-length patient-level habitat feature representation, followed by feature reduction (e). Intratumoral, habitat-based, and combined models were developed to predict 3-year cancer recurrence. The combined model integrated intratumoral and habitat radiomic risk scores using a stacked XGBoost framework to stratify patients into high- and low-risk recurrence groups (f).

**Figure 2.**
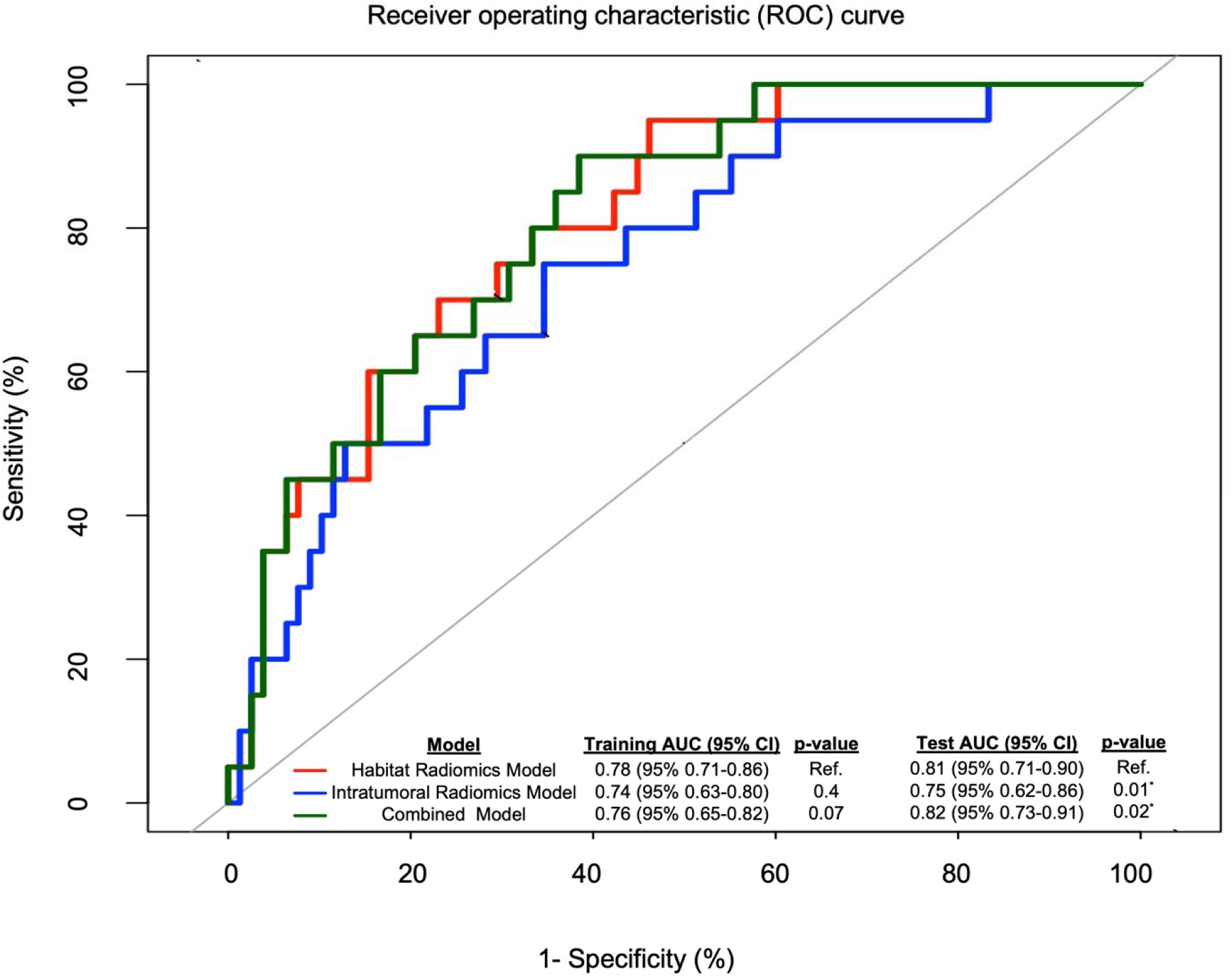
Receiver operating characteristic (ROC) curves comparing intratumoral, habitat, and combined models for predicting 3-year recurrence in the test cohort. Test-set ROC curves are shown; training AUC values are provided for reference. Differences in AUC were assessed using DeLong’s test, with the habitat-based model used as the reference.

Clinical and demographic variables were obtained the institutional Cancer Registry. Missing clinical data were imputed using class-specific medians and processed separately within the training and test cohorts. Disease recurrence was defined as the occurrence of local, regional, or distant tumor progression following surgical resection. This study was approved by an institutional review board (Advarra Inc)

### CT Image Processing

The CT images were standard-of-care imaging studies performed on scanners from Siemens and GE Medical Systems, with slice thickness ranging from 1 to 5 mm and in-plane spatial resolution between 0.55 and 0.98 mm. All images were reconstructed on a 512 × 512 matrix, and all patient identifiers were removed prior to analysis.

Intratumoral segmentation was generated by an imaging scientist with 14 years of experience using a single-click ensemble segmentation approach using Health-Myne software [25]. Segmentations were randomly reviewed in approximately 10% of cases by a radiologist with 15 years of experience for quality assurance purposes. All final tumor segmentations were stored in RTSTRUCT (Radiotherapy Structure Set) format for subsequent analysis.

CT images and corresponding region-of-interest (ROI) segmentations were resampled to an isotropic voxel resolution of 1 mm × 1 mm × 1 mm to ensure spatial consistency across patients. Cubic interpolation was applied to the CT images, while nearest-neighbor interpolation was used for the segmentation masks to preserve label integrity. All image preprocessing steps were implemented in MATLAB (version 2024b).

### Generation of Habitat Maps and Habitat Radiomic Features

Tumor habitats were generated using a two-level clustering framework consisting of individual-level supervoxel segmentation followed by population-level clustering. At the individual level, resampled CT images were min-max normalized within the tumor region. To capture local textural heterogeneity within the tumor, a slice-wise two-dimensional entropy filter was applied. Then, normalized CT images and entropy masks were fused to form the input for supervoxel segmentation.

Supervoxels were generated using a three-dimensional Simple Linear Iterative Clustering (SLIC) algorithm [26], with the number of supervoxels adaptively determined based on tumor size to account for inter-patient variability in tumor volume. For each supervoxel, histogram-based intensity and entropy features were computed to characterize supervoxel-level phenotypes and pooled across all patients for subsequent population-level clustering.

At the population level, unsupervised k-means clustering was applied to the pooled supervoxel feature set. The number of habitats (K = 3) was determined using consensus clustering–based stability analysis [27]. Cluster labels were defined at the population level and then mapped back to each patient to generate voxel-wise habitat maps. This two-level clustering strategy enables the characterization of spatially distinct tumor subregions while preserving population-level phenotypic consistency. Additional methodological details of habitat generation are provided in the Supplementary Data.

Habitat labels were used to define region of interest masks and radiomic features were extracted separately for each habitat within the tumor volume using the PyRadiomics library [28]. Feature extraction parameters included intensity normalization (scale = 1) and removal of outliers beyond three standard deviations. Gray-level discretization was performed using a fixed bin width of 25 Hounsfield units.

For each habitat, a total of 107 radiomic features were extracted, including 18 first-order statistics, 14 shape-based features, 70 gray-level texture features, and 5 neighboring gray tone difference features.

### Habitat Feature Aggregation Strategies

Due to inter-patient variability in tumor composition, not all patients exhibited all three habitats. Habitat assignment was driven by population-level clustering of similar supervoxels and was not constrained to occur uniformly across patients. As a result, some patients contained a single habitat (Habitat 1, Habitat 2, or Habitat 3), others had two habitats (Habitat 1+2, Habitat 1+3, or Habitat 2+3), and some contained all three habitats.

To enable consistent patient-level representation for downstream machine learning analyses, multiple feature aggregation strategies were implemented to summarize habitat-level radiomic features. These strategies included maximum, mean, minimum, variance, and sum of habitat-level feature values. Following feature aggregation, a fixed-dimensional feature vector consisting of 107 features was obtained for each patient, regardless of the number of habitats present. Similar habitat-level feature aggregation strategies have been reported previously [29]. Each aggregation strategy was evaluated using an identical feature selection pipeline and the same classifier (ridge-regularized logistic regression) within a nested cross-validation framework applied to the training set to ensure a fair comparison. The final models were then evaluated on an independent, test set. Separate models were trained and evaluated on both training and test sets for each strategy to identify the optimal aggregation approach, as described in the Model Development section, and the results are summarized in Table 2. Calculation details of these aggregation strategies are provided in the Supplementary Data.

### Intratumoral Radiomic Feature Calculation

The whole-tumor segmentation was used to define the region-of-interest, and radiomic features were extracted from the entire tumor volume using the same PyRadiomics [28] settings as those applied in the habitat-based radiomics described above. A total of 107 radiomic features were extracted, including 18 first-order statistics, 14 shape-based features, 70 gray-level texture features, and 5 neighboring gray tone difference features.

### Statistical Analyses

To compare demographic and clinical variables between the training and test cohorts, we utilized Fisher’s exact test for categorical variables and the Wilcoxon rank-sum test for continuous variables. All analyses were conducted using R software (version 4.4.2) [30]. Statistical significance was defined as a two-sided p-value < 0.05.

Min-max normalization (−1 to 1) was applied to scale all radiomic features and a multistep strategy was applied to reduce number of intratumoral radiomic features (N = 107) and to the aggregated habitat-based radiomic features (N = 107). Feature reduction was performed on the training set to prevent information leakage. For habitat-based radiomics, feature selection was performed separately for each feature aggregation strategy. To mitigate feature redundancy, highly correlated features (absolute Pearson correlation |r| > 0.90) and near-zero variance features were removed to limit multicollinearity and exclude non-informative features. Second, univariable analyses were performed using the Wilcoxon rank-sum test, and features with p-values < 0.20 were retained to remove clearly uninformative features while keeping potentially predictive candidates for multivariable selection. Next, the remaining features were included into LASSO logistic regression with 5-fold cross-validation, where the regularization parameter (λ) was selected to optimize the area under the ROC curve (AUC) [31] and yield a parsimonious and predictive feature set. Feature stability was assessed by repeating the full selection pipeline using bootstrap resampling (1000 iterations) on the training set only [27]. Selection frequency was computed for each feature across bootstrap iterations, and features selected in ≥70% of resamples were retained to prioritize stable and reproducible predictors, consistent with stability selection principles [27]. This procedure yielded the final feature sets for both the intratumoral and habitat-based models. The complete workflow is illustrated in Supplementary Figure S2.

Three radiomic classifiers were developed to predict 3-year recurrence as a binary outcome: (1) intratumoral radiomics only, (2) habitat radiomics only, and (3) combination of intratumoral and habitat radiomics. The intratumoral and habitat classifiers were constructed using ridge-regularized logistic regression (glmnet, α = 0) [32], with hyperparameter tuning performed via 7-fold nested cross-validation on the training cohort to predict recurrence probabilities. Seven-fold nested cross-validation was used to balance bias and variance while maintaining sufficient sample size within each fold. Final ridge logistic regression classifiers were fitted using the optimal regularization parameters (λ), determined independently for the habitat radiomics and intratumoral radiomics models. After hyperparameter tuning, the final classifiers were refit on the full training cohort using the selected optimal regularization parameter. This cross-validation procedure was used solely for model training and hyperparameter tuning and was distinct from the cross-validation steps applied during feature selection. The combination classifier was developed using a stacked learning framework [33,34], in which intratumoral- and habitat-based risk scores were integrated using an XGBoost meta-learner [35]. This approach enabled integration of intratumoral and habitat radiomics information for recurrence prediction. The three final classifiers were applied to predict 3-year recurrence in the unseen test cohort. Model discrimination was quantified using the area under the receiver operating characteristic curve (AUC). Differences in AUC between models were assessed using DeLong’s test. Model interpretability for 3-year recurrence prediction was evaluated using SHapley Additive exPlanations (SHAP) analysis [36].

For each classifier, the probability of recurrence was calculated for patient in the training set and then distribution- (median and tertiles) and machine learning–based (Classification and Regression Tree [CART] adapted for survival time, k-means clustering, and Gaussian mixture models [GMM]) approaches were utilized to identify the most informative threshold to stratify patients into low- and high-risk groups of recurrence. Survival analyses comparing the high-risk and low-risk groups were performed using Cox regression, Kaplan-Meier survival estimates, and log-rank test. Recurrence-free survival was the outcome variable. Absolute difference in 3-year recurrence-free survival probability between the high-risk and low-risk groups (Delta, Δ) was calculated. The thresholds were applied to the test dataset to assess model validation. Additional, details of these analyses are provided in the Supplementary Data.

## Results

### Patient Characteristics

There were no statistically significant differences between the training and test datasets for the demographic and clinical characteristics (Table 1). Three-year recurrence was observed in 21% of the training dataset and 20.4% of the test dataset.

**Table 1.** Comparison of Patient Characteristics Between Training and Test Cohorts.

|  |  |  |  |
| --- | --- | --- | --- |
| Patient Characteristics | Training Cohort N = 195 | Test Cohort N = 98 | p - value |
| Mean Age ± SD | 67.4 (10.2) | 67.6 (9.5) | 0.88 |
| Sex, N (%) |  |  | 0.81 |
| Female | 101 (51.8) | 53 (54.1) |  |
| Male | 94 (48.2) | 45 (45.9) |  |
| Race, N (%) |  |  | 0.43 |
| White | 181 (92.8) | 94 (95.9) |  |
| Other | 14 (7.2) | 4 (4.1) |  |
| Smoking History, N (%) |  |  | 0.62 |
| Never | 21 (10.8) | 8 (8.2) |  |
| Current/Former | 174 (89.2) | 90 (91.8) |  |
| TNM Stage, N (%) |  |  | 0.41 |
| 1A | 70 (35.9) | 42 (42.9) |  |
| 1B | 36 (18.5) | 20 (20.4) |  |
| 2A | 15 (7.7) | 10 (10.2) |  |
| 2B | 32 (16.4) | 11 (11.2) |  |
| 3A | 42 (21.5) | 15 (15.3) |  |
| Surgery Type, N (%) |  |  | 0.35 |
| Lobectomy | 126 (64.6) | 73 (74.5) |  |
| Segmentectomy | 38 (19.5) | 13 (13.3) |  |
| VATS | 10 (5.1) | 5 (5.1) |  |
| Other | 21 (10.8) | 7 (7.1) |  |
| Adjuvant Therapy, N (%) |  |  | 0.42 |
| None | 121 (62.1) | 67 (68.4) |  |
| Chemotherapy | 49 (25.1) | 23 (23.5) |  |
| Radiation or Chemoradiation | 25 (12.8) | 8 (8.2) |  |
| Tumor Site, N (%) |  |  | 0.81 |
| Main Bronchus or Overlapping Lesion | 11 (5.6) | 4 (4.1) |  |
| Upper Lobe | 111 (56.9) | 52 (53.1) |  |
| Middle Lobe | 5 (2.6) | 3 (3.1) |  |
| Lower Lobe | 68 (34.9) | 39 (39.8) |  |
| Histology, N (%) |  |  | 0.81 |
| SCC | 63 (32.3) | 28 (28.6) |  |
| AC | 123 (63.1) | 65 (66.3) |  |
| Other | 9 (4.6) | 5 (5.1) |  |
| Regional LN Metastases, N (%) |  |  | 0.69 |
| No | 145 (74.4) | 70 (71.4) |  |
| Yes | 50 (25.6) | 28 (28.6) |  |
| 3-Year Recurrence, N (%) |  |  | 0.90 |
| No | 154 (79.0) | 78 (79.6) |  |
| Yes | 41 (21.0) | 20 (20.4) |  |
| AC, adenocarcinoma; LN, lymph node; SCC, squamous cell carcinoma; SD, standard deviation; TNM, tumor, node, metastases; VATS, video-assisted thoracoscopic surgery. |  |  |  |

**Table 2.**
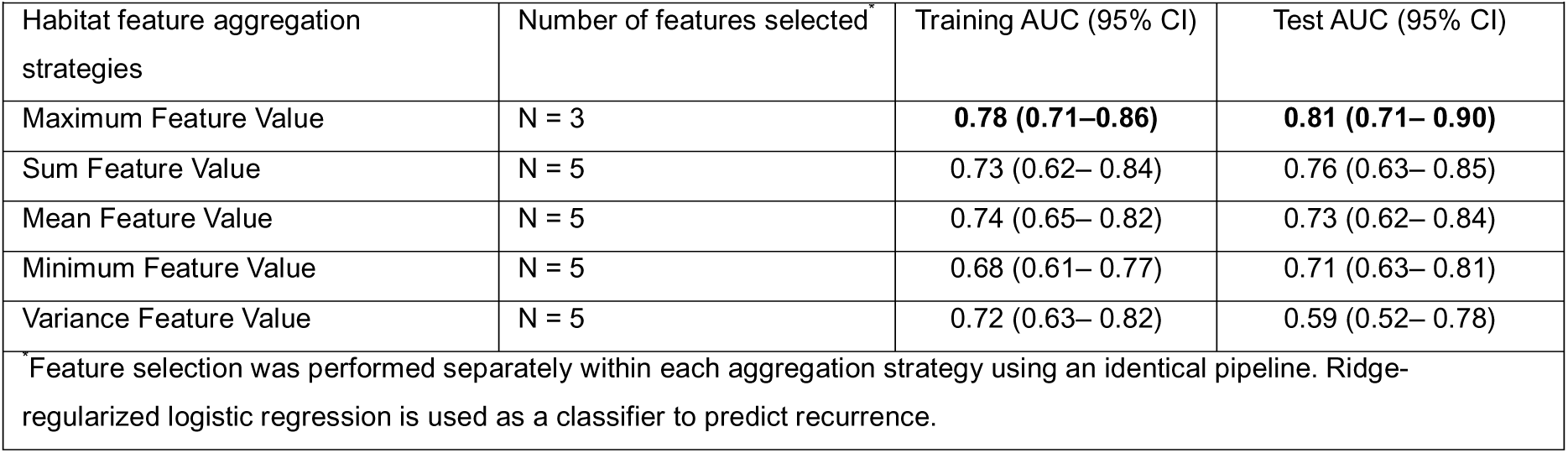
Performance Comparison of Different Habitat Aggregation Strategies.

|  |  |  |  |
| --- | --- | --- | --- |
| Habitat feature aggregation strategies | Number of features selected<sup>*</sup> | Training AUC (95% CI) | Test AUC (95% CI) |
| Maximum Feature Value | N = 3 | **0.78 (0.71–0.86)** | **0.81 (0.71– 0.90)** |
| Sum Feature Value | N = 5 | 0.73 (0.62– 0.84) | 0.76 (0.63– 0.85) |
| Mean Feature Value | N = 5 | 0.74 (0.65– 0.82) | 0.73 (0.62– 0.84) |
| Minimum Feature Value | N = 5 | 0.68 (0.61– 0.77) | 0.71 (0.63– 0.81) |
| Variance Feature Value | N = 5 | 0.72 (0.63– 0.82) | 0.59 (0.52– 0.78) |
| <sup>*</sup> Feature selection was performed separately within each aggregation strategy using an identical pipeline. Ridge-regularized logistic regression is used as a classifier to predict recurrence. |  |  |  |

### Radiomic models to predict recurrence

Radiomic models were developed to predict 3-year recurrence as a binary outcome. The intratumoral radiomic model, which was comprised of three features (Surface Volume Ratio, Elongation, and Short Run High Gray Level Emphasis), yielded AUCs of 0.74 (95% 0.63-0.80) and 0.75 (95% 0.62-0.86) in the training and test sets, respectively. All aggregation strategies were systematically compared (Table 2), and the maximum-value aggregation yielded the highest AUCs on both the training and test sets compared to other approaches (sum-value, mean-value, minimum-value, and variance-value aggregation strategies). Accordingly, the habitat radiomics model, hereafter defined using maximum-value aggregation, achieved AUCs of 0.78 (95% 0.71–0.86) and 0.81 (95% 0.71–0.90) in the training and test sets, respectively. The habitat radiomics model was comprised of three features: Large Dependence Emphasis, Large Dependence High Gray Level Emphasis, and Major Axis Length. The combined model concatenated the intratumoral and habitat radiomics models and yielded AUCs of 0.76 (95% 0.65-0.82) and 0.82 (95% 0.73-0.91) in the training and test sets, respectively. Based on the DeLong’s test, both the combined model (p = 0.02) and the intratumoral radiomics model (p = 0.01) showed statistically significant differences in test-set performance compared with the habitat radiomics model.

### Recurrence-free survival

Table 3 provides risk estimates and performance statistics for the models by training and test datasets; the 3-year survival rates for the low- and high-risk patients and the delta between the low- and high-risk populations are also presented. Based on the radiomic models to predict recurrence, patients classified as high-risk based on the CART probability of recurrence were consistently associated with statistically significantly inferior survival outcomes for all three models in both the training and test cohorts (Figures 3g, h, j). This pattern yielded larger hazard ratios and a larger delta for percentage of 3 year recurrence between risk groups compared to other approaches (Table 3). In the test cohort, the combined model yielded the largest HR (HR = 8.43; 95% CI 2.47 – 28.81) compared to the habitat radiomics only (HR = 5.41; 95% CI 2.08 – 14.09) and intratumoral radiomics only (HR = 3.54; 95% CI 1.45 – 8.66) models. The deltas between the high- and low-risk cohorts for the 3-year survival rates was 33% for the combined model and 32% and 25% for the habitat radiomics only and intratumoral radiomics only models, respectively.

**Figure 3.**
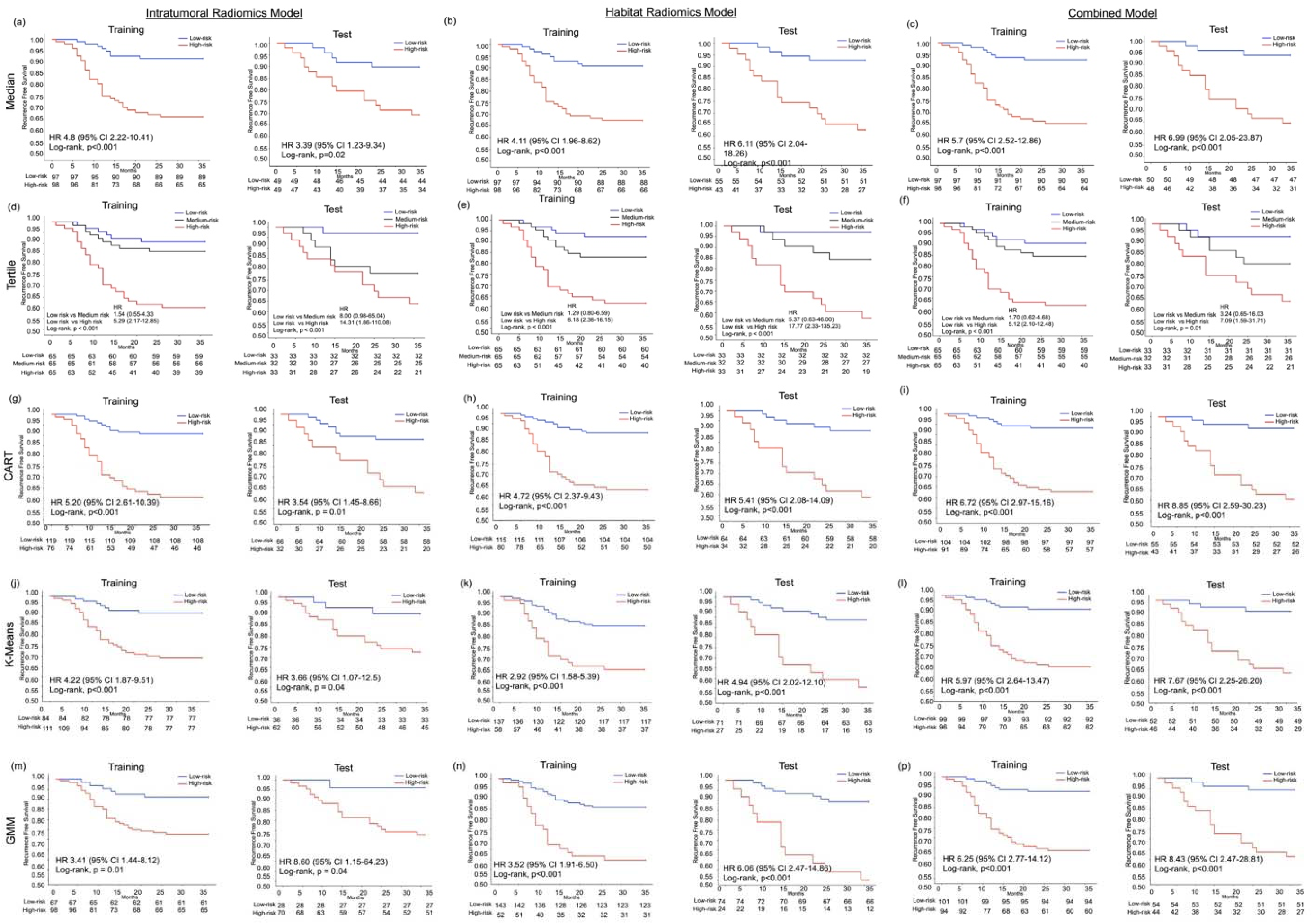
Kaplan–Meier curves illustrating 3-year recurrence-free survival for high-risk and low-risk groups stratified by the intratumoral radiomics model, habitat radiomics model, and the combined model (intratumoral + habitat) across different thresholding strategies, including median (a,b,c), tertile (d,e,f), classification and regression tree (CART)–based (g,h,i), k-means (j,k,l), and Gaussian mixture model (GMM)–based approaches (m,n,p), in the training and test cohorts. Survival differences between risk groups were assessed using the log-rank test, and hazard ratios (HRs) were estimated using univariable Cox proportional hazards regression.

**Table 3.** Performance Evaluation of All Models in the Training and Test Cohorts.

| Threshold strategies | Datasets | Model | HR (95% CI) | Log-rank p-value | C-index (95% CI) | 3-year % survival rate for low risk | 3-year % survival rate for high risk | Delta of 3-year % survival rate |
| --- | --- | --- | --- | --- | --- | --- | --- | --- |
| Median | Training | Habitat | 4.11 (1.96 – 8.62) | < 0.001<sup>+</sup> | 0.70 (0.63 – 0.78) | 91% | 67% | 24% |
|  |  | Intratumoral | 4.80 (2.22– 10.41) | < 0.001<sup>+</sup> | 0.71 (0.63 – 0.79) | 92% | 66% | 26% |
|  |  | Combined | 5.70 (2.52 –12.86) | < 0.001<sup>+</sup> | 0.72 (0.64 – 0.79) | 93% | 65% | 28% |
|  | Test | Habitat | 6.10 (2.04 – 18.26) | < 0.001<sup>+</sup> | 0.75 (0.66 – 0.84) | 93% | 63% | 30% |
|  |  | Intratumoral | 3.39 (1.23 – 9.34) | 0.02<sup>+</sup> | 0.72 (0.61 – 0.83) | 90% | 69% | 21% |
|  |  | Combined | 6.99 (2.05 – 23.87) | < 0.001<sup>+</sup> | 0.77 (0.67 – 0.86) | 94% | 65% | 29% |
| Tertile | Training | Habitat (Low vs Medium) | 2.29 (0.80 – 6.59) | < 0.001<sup>+</sup> | 0.69 (0.62 – 0.76) | 92% | 83% | 9% |
|  |  | Habitat (Low vs High) | 6.18 (2.36 – 16.15) |  |  | 92% | 62% | 30% |
|  |  | Intratumoral Low vs Medium) | 1.54 (0.55 – 4.33) | < 0.001<sup>+</sup> | 0.69 (0.61 – 0.76) | 91% | 86% | 5% |
|  |  | Intratumoral (Low vs High) | 5.29 (2.17–12.85) |  |  | 91% | 60% | 31% |
|  |  | Combined (Low vs Medium) | 1.70 (0.62 – 4.68) | < 0.001<sup>+</sup> | 0.68 (0.61 – 0.76) | 91% | 85% | 6% |
|  |  | Combined (Low vs High) | 5.12 (2.10 – 12.48) |  |  | 91% | 62% | 29% |
|  | Test | Habitat (Low vs Medium) | 5.37 (0.63 – 46.00) | < 0.001<sup>+</sup> | 0.75 (0.66 – 0.84) | 97% | 84% | 13% |
|  |  | Habitat (Low vs High) | 17.77 (2.33 – 135.23) |  |  | 97% | 58% | 39% |
|  |  | Intratumoral (Low vs Medium) | 8.00 (0.98 – 65.04) | < 0.001<sup>+</sup> | 0.70 (0.61 – 0.80) | 97% | 78% | 19% |
|  |  | Intratumoral (Low vs High) | 14.31 (1.86 – 110.08) |  |  | 97% | 64% | 33% |
|  |  | Combined (Low vs Medium) | 3.24 (0.65 – 16.03) | 0.01<sup>+</sup> | 0.69 (0.58 – 0.79) | 94% | 81% | 13% |
|  |  | Combined (Low vs High) | 7.09 (1.59 – 31.71) |  |  | 94% | 64% | 30% |
| CART | Training | Habitat | 4.72 (2.37 – 9.43) | < 0.001<sup>+</sup> | 0.70 (0.63 – 0.78) | 90% | 63% | 27% |
|  |  | Intratumoral | 5.20 (2.61 – 10.39) | < 0.001<sup>+</sup> | 0.71 (0.63 – 0.79) | 91% | 61% | 30% |
|  |  | Combined | 6.72 (2.97 – 15.16) | < 0.001<sup>+</sup> | 0.72 (0.64 – 0.79) | 93% | 63% | 30% |
|  | Test | Habitat | 5.41 (2.08 – 14.09) | < 0.001<sup>+</sup> | 0.78 (0.69 – 0.87) | 91% | 59% | 32% |
|  |  | Intratumoral | 3.54 (1.45 – 8.66) | 0.01<sup>+</sup> | 0.72 (0.61 – 0.83) | 88% | 63% | 25% |
|  |  | Combined | 8.43 (2.47 – 28.81) | < 0.001<sup>+</sup> | 0.77 (0.67 – 0.86) | 94% | 61% | 33% |
| K-means | Training | Habitat | 2.92 (1.58 – 5.39) | < 0.001<sup>+</sup> | 0.70 (0.63 – 0.78) | 85% | 64% | 21% |
|  |  | Intratumoral | 3.50 (1.47 – 8.32) | < 0.001<sup>+</sup> | 0.71 (0.63 – 0.79) | 91% | 72% | 19% |
|  |  | Combined | 7.32 (2.14 – 25.01) | < 0.001<sup>+</sup> | 0.77 (0.67 – 0.86) | 94% | 64% | 30% |
|  | Test | Habitat | 4.94 (2.02 – 12.10) | < 0.001<sup>+</sup> | 0.78 (0.69 – 0.87) | 89% | 56% | 33% |
|  |  | Intratumoral | 9.54 (1.28 – 71.28) | 0.03<sup>+</sup> | 0.72 (0.61 – 0.83) | 97% | 72% | 25% |
|  |  | Combined | 5.97 (2.64 –13.47) | < 0.001<sup>+</sup> | 0.72 (0.64 – 0.79) | 93% | 65% | 28% |
| GMM | Training | Habitat | 3.52 (1.91 – 6,5) | < 0.001<sup>+</sup> | 0.70 (0.63 – 0,78) | 86% | 60% | 26% |
|  |  | Intratumoral | 3.41 (1.44 – 8.12) | 0.01<sup>+</sup> | 0.71 (0.63 – 0.79) | 91% | 73% | 18% |
|  |  | Combined | 6.25 (2.77 – 14.12) | < 0.001<sup>+</sup> | 0.72 (0.64 – 0.79) | 93% | 64% | 29% |
|  | Test | Habitat | 6.06 (2.47 – 14.86) | < 0.001<sup>+</sup> | 0.78 (0.69 – 0.87) | 89% | 50% | 39% |
|  |  | Intratumoral | 8.60 (1.15 – 64.23) | 0.04<sup>+</sup> | 0.72 (0.61 – 0.83) | 96% | 73% | 23% |
|  |  | Combined | 8.04 (2.35 – 27.47) | < 0.001<sup>+</sup> | 0.77 (0.67 – 0.86) | 94% | 62% | 32% |

HR: Hazard Ratios; CI: Confidence interval.; GMM: Gaussian Mixture Model; CART: Classification and Regression Trees. Delta represents the absolute difference in 3-year survival probability between the low-risk and high-risk groups. For tertile-based analyses, log-rank p-values were derived from overall three-
group comparisons, while hazard ratios are reported for pairwise comparisons. The C-index reflects overall model discrimination.

Patient-level characteristic between high-risk and low-risk recurrence groups for the intratumoral, habitat, and combined models are presented in Supplementary Table 1 (training cohort) and Supplementary Table 2 (test cohort). Consistent findings were observed across alternative thresholding strategies, including median-, tertile-, K-means, and GMM approaches, with similar patterns of risk separation and statistically significant differences in recurrence-free survival (Figures 3a to 3p; Table 3).

### Model Interpretability and Feature Importance by SHAP Analysis

Figure 4 presents SHAP beeswarm plots (Fig 4 a, c, e) and SHAP feature importance plots (Fig 4 b, d, f) for the intratumoral radiomics, habitat radiomics, and combined models. The SHAP beeswarm plots summarize the global impact of each feature on model predictions across all patients, while the SHAP feature importance plots summarize the relative contribution of each feature within its respective model. For the intratumoral radiomics model, the most influential features, ranked by mean absolute SHAP values, were Surface Volume Ratio, Elongation, and Short Run High Gray Level Emphasis (Figure 4a and 4b). For the habitat radiomics model, the top contributing features were Large Dependence Emphasis, Large Dependence High Gray Level Emphasis, and Major Axis Length (Figure 4c and 4d). For the combined model, the habitat-based risk score contributed more strongly to recurrence prediction than the intratumoral risk score, with higher mean absolute SHAP values (0.64 vs. 0.36, respectively), indicating a greater influence of habitat-derived information on the final prediction (Figure 4e and 4f).

**Figure 4.**
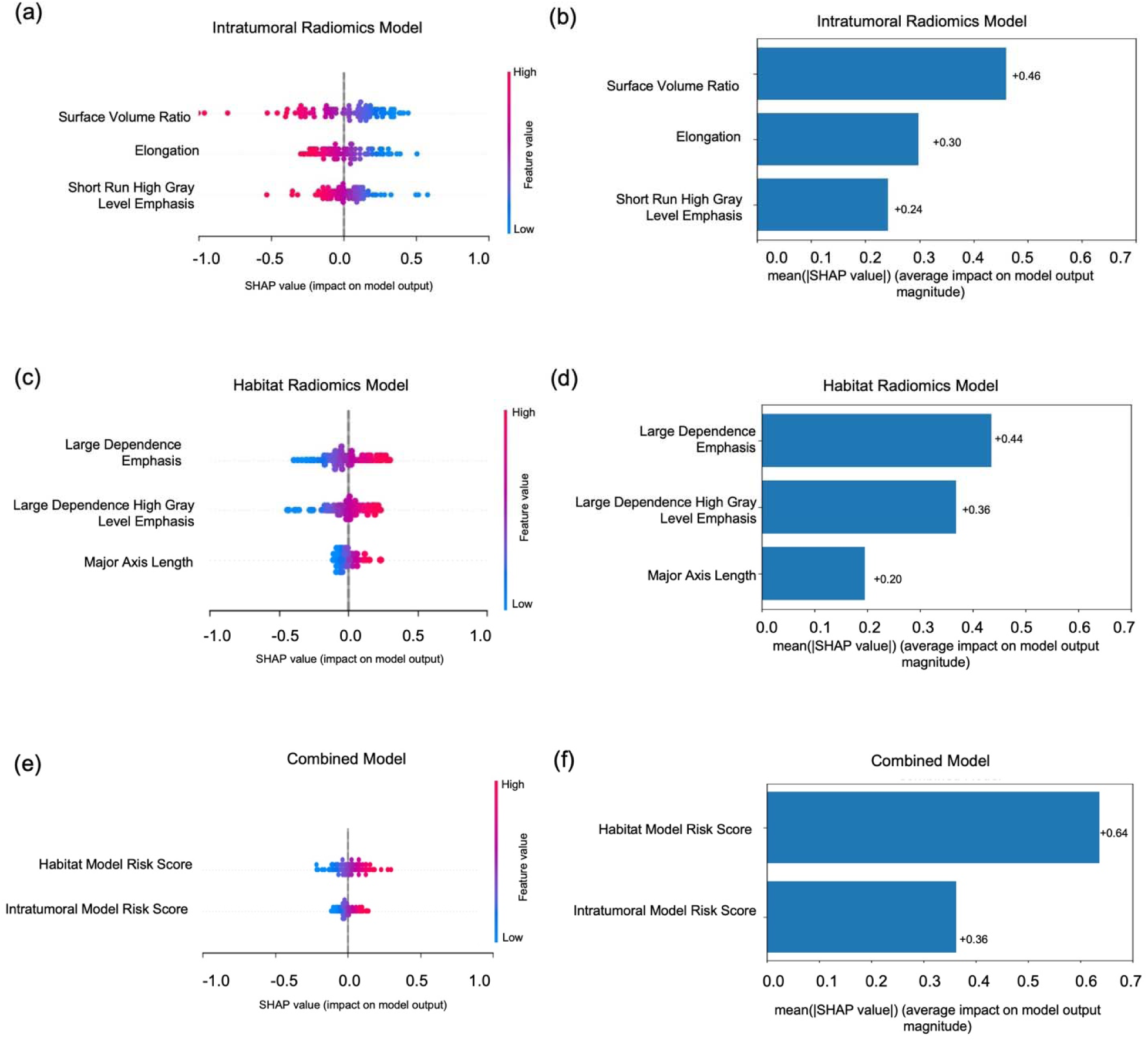
**SHAP-based feature importance analysis of intratumoral, habitat, and combined radiomics models**. SHAP beeswarm and summary bar plots were used to evaluate feature importance and model interpretability. (a, c, e) Beeswarm plots illustrate the distribution of SHAP values for selected features in the intratumoral, habitat, and combined models, respectively. Each point represents an individual patient and is colored according to the corresponding feature value (blue: low, red: high). The x-axis indicates the impact of each feature on the model output, with positive SHAP values corresponding to a higher predicted risk of recurrence and negative values corresponding to a lower predicted risk. (b, d, f) Bar plots show the mean absolute SHAP values, summarizing the relative contribution of each feature within its respective model.

## Discussion

In this study, we conducted deployed a rigorous and systematic approach to identify the most informative radiomic models to predict disease recurrence among NSCLC patients who received surgical resection as first line of treatment. Multiple feature aggregation strategies for habitat-based radiomics features were compared, habitat and intratumor radiomic features were evaluated to identify the most informative classifiers to predict 3-year recurrence, and then distributional and machine learning approaches were deployed to identify optimal thresholds to discriminate between high- and low-risk patients in time-dependent analyses. Overall, habitat-based radiomics exhibited more superior performance than intratumoral radiomics only; however, combining habitat- and intratumoral-derived features further improved model performance. Across multiple habitat feature aggregation strategies, the maximum-value aggregation approach yielded the most favorable performance. In addition, risk stratification results were consistent across different thresholding strategies, indicating that model performance was not driven by a specific cut-point selection method. To the best of our knowledge, this study represents the first systematic evaluation of habitat-based radiomics for recurrence risk stratification in NSCLC.

Habitat-based radiomics partitions a region-of-interest (e.g., tumor) into spatially defined subregions according to differences in imaging characteristics [37]. Since radiomic features reflect the underlying pathophysiology of a region-of-interest [17,21], habitat radiomics likely represent biologically distinct tumor microenvironments. To that end, our analyses revealed that the habitat radiomics model was superior to intratumoral radiomics model which is consistent with other studies [21,29,38]. Thus, the additional biological underpinnings captured by habitat radiomics yield improved statistical performance over intratumoral radiomics. Moreover, our systematic approach to model development revealed that the maximum-value habitat feature aggregation strategy yielded superior performance compared to the aggregation strategies. From a modeling perspective, such aggregation is also necessary to obtain a consistent patient-level representation suitable for downstream classical machine learning analyses, given the variable number of habitats observed across patients. While several studies have explored maximum-value aggregation or k-nearest neighbor–based imputation to address this issue [21,29,39–41], many habitat radiomics workflows do not explicitly report how habitat-level features are integrated into patient-level representations for downstream prediction. Notably, the maximum-value habitat feature aggregation strategy also demonstrated superior performance in our prior habitat-based radiomics study, albeit for a different clinical endpoint [29]. Similar findings have been reported by Han et al. [39], who compared mean-based, maximum-value–based, and k-nearest neighbor imputation strategies for integrating heterogeneous tumor subregions into patient-level representations for machine learning–based prediction, and observed that maximum-value aggregation yielded the most optimal predictive performance. The consistent selection of this aggregation approach across independent studies and outcomes suggests that maximum-value aggregation may represent a reproducible strategy for summarizing habitat-level radiomic information. Additionally, habitat radiomics has been widely applied in recent literature across cancer types and imaging modalities, supporting the use of subregion-based phenotyping to reflect biologically distinct tumor microenvironments and prognostic relevance [42].

In the habitat radiomics model, three radiomic features (Large Dependence Emphasis, Large Dependence High Gray Level Emphasis, and Major Axis Length) were identified as the most informative to predict recurrence. Among these, Large Dependence Emphasis, which characterizes spatially connected regions with similar gray-level intensities [43,44], was identified as the most informative feature. Higher values of this feature, reflecting more homogeneous and solid intratumoral habitats, were associated with an increased likelihood of recurrence. This finding is consistent with our group’s prior work [24], in which the same feature was identified as a prognostic marker at the intratumoral level, and now extends its relevance to habitat-based characterization of tumor heterogeneity. Although Large Dependence Emphasis has been widely used as an intratumoral radiomic feature, to the best of our knowledge, this is the first study to report its prognostic value within a habitat-based radiomics framework for lung cancer. Similarly, Zheng *et al.* [45] reported Large Dependence High Gray Level Emphasis as a significant habitat radiomic feature for predicting recurrence in lung adenocarcinoma, supporting the broader relevance of GLDM-based dependence features. In addition, higher values of Major Axis Length were associated with an increased likelihood of recurrence, consistent with prior radiological studies showing that greater tumor extent is associated with worse clinical outcomes in lung cancer [46]. Notably, Major Axis Length identified as a key habitat-level feature in this study, was also reported in our prior habitat radiomics study for HPV prediction [29], suggesting that tumor extent–related habitat morphology may represent a reproducible imaging signature across distinct clinical endpoints. Our results extend this body of work by demonstrating that spatial intratumoral heterogeneity captured from preoperative CT alone provides meaningful prognostic information for recurrence risk stratification in surgically resected NSCLC.

For the intratumoral radiomics model, lower values of Surface Volume Ratio, Elongation, and Short Run High Gray Level Emphasis contribute to higher likelihood of recurrence, indicating that global tumor morphology and fine-scale texture patterns are informative. Surface Volume Ratio and Elongation describe overall tumor geometry, where lower values reflect a more compact morphology. Prior radiomics studies have demonstrated that shape-based features are associated with tumor aggressiveness and survival across multiple cancer types, including head and neck, lung, and glioma cancers [47–49]. Additionally, lower value of Short Run High Gray Level Emphasis, a texture feature capturing fine-scale high-intensity patterns, was associated with higher recurrence risk. Texture heterogeneity metrics have consistently shown prognostic relevance and have been linked to tumor biology, treatment response, and survival in radiomics studies [47,50,51].

The combined model integrates intratumoral and habitat-based risk scores using a stacked learning approach, allowing complementary tumor information to be jointly leveraged. In this model, the habitat-based risk score contributed more strongly than the intratumoral risk score, highlighting the added prognostic value of habitat-level information beyond whole-tumor features. These findings suggest that spatial tumor heterogeneity relevant to recurrence risk is more effectively captured through habitat-based analysis. Importantly, the combined model preserves established whole-tumor information while augmenting it with habitat-level heterogeneity, which may enhance clinical interpretability and acceptance. The combined model demonstrated consistent prognostic performance across multiple threshold strategies, supporting the presence of a clinically meaningful underlying risk signal. By integrating intratumoral and habitat-based radiomics, this approach captures complementary tumor characteristics, with habitat features providing additional insight into spatial heterogeneity. While current clinical recommendations rely primarily on TNM staging to guide postoperative management and adjuvant treatment decisions [52,53], radiomic modeling—particularly when incorporating habitat-based features—may provide additional, nonredundant information to identify patients at higher risk of recurrence within the same stage. Importantly, this framework can be applied preoperatively using routine contrast-enhanced CT imaging, without the need for invasive procedures or tissue sampling, highlighting its potential as an accessible and early prognostic tool to support personalized risk stratification and surveillance planning.

While we acknowledge there are some limitations of this study including the modest sample size and all patients were derived from single institution which may limit generalizability. However, the rigorous and systematic approach deployed can be utilized as a framework to validate our observations in larger and more diverse patient cohorts. Nonetheless, this study revealed that habitat-based radiomics provided superior statistically significant improvement over intratumoral radiomics for predicting recurrence in NSCLC. Moreover, combining habitat- and intratumoral-radiomics yielded a modest increase in performance compared to habitat only and intratumoral models. These findings highlight the value of capturing intratumoral heterogeneity using preoperative CT imaging and support the potential role of habitat-based radiomics as a noninvasive tool to aid personalized postoperative risk assessment and surveillance planning.

## Data Availability

The data underlying this article will be made available upon reasonable request.

## Acknowledgements

This work has been supported in part by the Collaborative Data Services Core (CDSC) and Quantitative Imaging Core (QIC) Core at the H. Lee Moffitt Cancer Center & Research Institute, an NCI designated Comprehensive Cancer Center (P30 CA076292).

## Data Availability

The data underlying this article will be made available upon reasonable request.

## Dedication

This manuscript is dedicated to the memory of Dr. Robert J. Gillies (1953-2022).

