## Supplementary Material for "Preoperative CT-Based Habitat Radiomics Classifiers Predict Recurrence in Non-Small Cell Lung Cancer"

**Supplementary Data**

**1. Mathematical Framework of the Two-Level Clustering Approach for Habitat Generation**

This section provides a detailed mathematical and technical description of the two-level clustering framework used to derive tumor habitats from radiomic features.

**1. 1. Individual-Level Clustering (Supervoxel Generation)**

At the patient level, each tumor region of interest (ROI) was processed to generate a fused image that integrates normalized CT intensity values with entropy-based texture information. This fusion was performed on a voxel-wise basis as follows:

Fused Image=CT_ROI_ + CT_LocalEntropy_

The resulting fused image was subsequently oversegmented within the tumor region using the Simple Linear Iterative Clustering (SLIC) algorithm. The number of generated supervoxels was adaptively adjusted according to tumor size to ensure appropriate spatial representation. All implementations were performed in MATLAB (version 2024b) using a compactness parameter of 0.01.

SLIC partitions an image into spatially contiguous and intensity-homogeneous regions (supervoxels in 3D) by jointly considering intensity similarity and spatial proximity. The distance metric D used to assign voxels to clusters is defined as:

$$D=\sqrt{\left( \frac{d_{c}}{m} \right)^{2}+\left( \frac{d_{s}}{S} \right)^{2}}$$

where:

- *d_c_* is is the Euclidean distance in intensity space
- *d_s_* is the Euclidean spatial distance in voxel coordinates.
- *S* is approximate size of each supervoxel (grid interval)
- *m* is compactness parameter controlling the balance between intensity and spatial proximity

***Radiomics Feature Extraction from Each Supervoxel***

For each supervoxel, first-order statistical features were computed based on voxel intensity distributions derived from both the original CT image and the entropy-filtered image. A total of 20 features were extracted (10 per modality), including: skewness, kurtosis, mean, median, first quartile, third quartile, interquartile range, standard deviation, variance, and energy. Each supervoxel was therefore represented as a 20-dimensional feature vector, capturing both intensity and texture heterogeneity.

**1.2. Population-Level Clustering (Habitat Generation)**

To identify shared imaging patterns across patients, all supervoxel-level feature vectors were aggregated into a single population-level dataset. The K-means clustering algorithm was then applied to this combined feature space to group supervoxels into distinct habitat regions. K-means is an unsupervised clustering technique that partitions data into K clusters by iteratively updating cluster centroids to minimize within-cluster variability. The optimization objective is defined as:

$$J=\sum_{i=1}^{N} \sum_{k=1}^{K} w_{ik}\times\parallel x_{i}-\mu_{k}\parallel^{2}$$

where:

- *J*: total clustering cost (also called the within-cluster sum of squared distances).
- *N*: number of data points (supervoxels).
- *K*: number of clusters (habitats).
- w_ik:_ indicator variable (1 if data point *i* belongs to cluster *k*, otherwise 0)
- x_i:_ feature vector of data point *i*.
- μ_k:_ centroid of cluster *k*.
- ∥x_i_-μ_k_ ∥^2^ : squared Euclidean distance between data point *i* and centroid *k*.

This two-level clustering framework enables the identification of spatially coherent and biologically meaningful tumor subregions that reflect intratumoral heterogeneity. All clustering procedures were implemented in MATLAB (version 2024b).

**Definition of ΔCDF**

ΔCDF (Delta Area Under the Cumulative Distribution Function) is a measure of cluster stability used in consensus clustering. For each number of clusters (K), a consensus matrix is generated through repeated subsampling and clustering. The cumulative distribution function (CDF) of the consensus values is then computed, and the area under the CDF curve (AreaCDF) is calculated.

ΔCDF is defined as the difference in AreaCDF between two consecutive cluster numbers:

ΔCDF(*K*) = AreaCDF(*K*) − AreaCDF(*K*−1)

A larger ΔCDF indicates a greater gain in clustering stability when moving from K−1 to K. As shown in Figure S1a, ΔCDF was evaluated across K values, and the largest increase was observed at K = 3. Based on this stability criterion, three habitats were selected for subsequent analyses.

The separation of the resulting habitat clusters is illustrated in Figure S1b, where supervoxel features are visualized in a principal component analysis (PCA) space. The three clusters show clear separation, supporting the selection of K = 3 as a stable and meaningful habitat solution.

**2.Habitat Feature Aggregation Strategies**

To determine which habitat-level feature set to use in the habitat radiomics classifier, we tested several feature aggregation strategies, including maximum feature value, sum, minimum, and variance. These strategies were designed to capture different aspects of the tumor subregions identified through habitat analysis. Among them, selecting features from the habitat with the maximum feature value yielded the best classification performance and was therefore used in the final habitat radiomics classifier. Each aggregation strategy was subsequently evaluated within a nested cross-validation framework, with a separate classifier trained for each strategy to identify the optimal aggregation approach based on cross-validated performance. Mathematical definitions of the aggregation strategies are provided below.

Let $f_{i,h}$ denote the value of radiomic feature $i$ in habitat $h$, and $H$ be the set of habitats present for a given patient. Aggregated feature values were computed as follows:

**Notation:**

- $i$: feature index (e.g., Sphericity, Entropy)
- $h$: habitat (e.g., H1, H2, H3)
- $f_{i,h}$: value of feature $i$ in habitat $h$

Maximum-value aggregation: $f_{i}^{\text{max}}$ represents the maximum value of feature $i$ across all habitats present in a patient.

$$f_{i}^{\text{max}}=\max_{h\in H}(f_{i,h})$$

Minimum-value aggregation: $f_{i}^{\text{min}}$ represents the minimum value of feature $i$ across all habitats present in a patient.

$$f_{i}^{\text{min}}=\min_{h\in H}(f_{i,h})$$

Mean-value aggregation: $f_{i}^{\text{mean}}$ represents the average value of feature $i$ across all habitats present in a patient.

$$f_{i}^{\text{mean}}=\frac{1}{\mid H\mid}\sum_{h\in H} f_{i,h}$$

Sum-value aggregation: $f_{i}^{\text{sum}}$ represents the sum of feature $i$ values across all habitats present in a patient.

$$f_{i}^{\text{sum}}=\sum_{h\in H} f_{i,h}$$

Variance-value aggregation: $f_{i}^{\text{var}}$ represents the variance of feature $i$ values across all habitats present in a patient.

$$f_{i}^{\text{var}}=\frac{1}{\mid H\mid}\sum_{h\in H} {(f_{i,h}-f_{i}^{\text{mean}})}^{2}$$

where $H$ includes only the habitats present for each patient.

**Supplemental Tables**

| **Supplementary Table 1.** Comparison of patient characteristics between high-risk and low-risk recurrence groups in the training cohort, defined using CART-derived cutpoints | | | | | | | | | |
| --- | --- | --- | --- | --- | --- | --- | --- | --- | --- |
| **Patient Characteristics** | **Habitat Radiomics Model** | | | **Intratumoral Radiomics Model** | | | **Combined Model** | | |
|  | **Low-risk**  **N = 115** | **High-risk**  **N = 80** | **p-value** | **Low-risk**  **N = 119** | **High-risk**  **N = 76** | **p-value** | **Low-risk**  **N = 104** | **High-risk**  **N = 91** | **p-value** |
| **Mean Age** ± **SD** | 67.10 (9.46) | 68.19 (9.66) | 0.44 | 67.17 (9.48) | 68.14 (9.64) | 0.49 | 67.43 (9.32) | 67.68 (9.82) | 0.86 |
| **Sex, N (%)**  Female  Male | 61 (53.0)  54 (47.0) | 40 (50.0)  40 (50.0) | 0.78 | 66 (55.5)  53 (44.5) | 35 (46.1)  41 (53.9) | 0.26 | 56 (53.8)  48 (46.2) | 45 (49.5)  46 (50.5) | 0.64 |
| **Race, N (%)**  White  Other | 109 (94.8)  6 (5.2) | 72 (90.0)  8 (10.0) | 0.32 | 111 (93.3)  8 (6.7) | 70 (92.1)  6 (7.9) | 0.98 | 99 (95.2)  5 (4.8) | 82 (90.1)  9 (9.9) | 0.27 |
| **Smoking History, N (%)**  Never  Current/Former | 12 (10.4)  103 (89.6) | 9 (11.2)  71 (88.8) | 0.99 | 13 (10.9)  106 (89.1) | 8 (10.5)  68 (89.5) | 0.99 | 11 (10.6)  93 (89.4) | 10 (11.0)  81 (89.0) | 0.99 |
| **TNM Stage, N (%)**  1A  1B  2A  2B  3A | 67 (58.3)  13 (11.3)  6 (5.2)  10 (8.7)  19 (16.5) | 3 (3.8)  23 (28.8)  9 (11.2)  22 (27.5)  23 (28.8) | <0.001* | 63 (52.9)  16 (13.4)  5 (4.2)  11 (9.2)  24 (20.2) | 7 (9.2)  20 (26.3)  10 (13.2)  21 (27.6)  18 (23.7) | <0.001* | 66 (63.5)  10 (9.6)  6 (5.8)  5 (4.8)  17 (16.3) | 4 (4.4)  26 (28.6)  9 (9.9)  27 (29.7)  25 (27.5) | <0.001* |
| **Surgery Type, N (%)**  Lobectomy  Segmentectomy  VATS  Other | 74 (64.3)  32 (27.8)  6 (5.2)  3 (2.6) | 52 (65.0)  6 (7.5)  4 (5.0)  18 (22.5) | <0.001* | 78 (65.5)  31 (26.1)  7 (5.9)  3 (2.5) | 48 (63.2)  7 (9.2)  3 (3.9)  18 (23.7) | <0.001* | 67 (64.4)  30 (28.8)  6 (5.8)  1 (1.0) | 59 (64.8)  8 (8.8)  4 (4.4)  20 (22.0) | <0.001* |
| **Adjuvant Therapy, N (%)**  None  Chemotherapy  Radiation or Chemoradiation | 84 (73.0)  21 (18.3)  10 (8.7) | 37 (46.2)  28 (35.0)  15 (18.8) | <0.001* | 88 (73.9)  19 (16.0)  12 (10.1) | 33 (43.4)  30 (39.5)  13 (17.1) | <0.001* | 83 (79.8)  12 (11.5)  9 (8.7) | 38 (41.8)  37 (40.7)  16 (17.6) | <0.001* |
| **Tumor Site, N (%)**  Main Bronchus or Overlapping Lesion  Upper Lobe  Middle Lobe  Lower Lobe | 4 (3.5)  65 (56.5)  3 (2.6)  43 (37.4) | 7 (8.8)  46 (57.5)  2 (2.5)  25 (31.2) | 0.42 | 3 (2.5)  75 (63.0)  3 (2.5)  38 (31.9) | 8 (10.5)  36 (47.4)  2 (2.6)  30 (39.5) | 0.05 | 3 (2.9)  62 (59.6)  3 (2.9)  36 (34.6) | 8 (8.8)  49 (53.8)  2 (2.2)  32 (35.2) | 0.34 |
| **Histology, N (%)**  SCC  AC  Other | 31 (27.0)  79 (68.7)  5 (4.3) | 32 (40.0)  44 (55.0)  4 (5.0) | 0.14 | 33 (27.7)  82 (68.9)  4 (3.4) | 30 (39.5)  41 (53.9)  5 (6.6) | 0.1 | 24 (23.1)  76 (73.1)  4 (3.8) | 39 (42.9)  47 (51.6)  5 (5.5) | 0.01* |
| **Regional LN Metastases, N (%)**  No  Yes | 96 (83.5)  19 (16.5) | 49 (61.2)  31 (38.8) | <0.001* | 94 (79.0)  25 (21.0) | 51 (67.1)  25 (32.9) | 0.1 | 88 (84.6)  16 (15.4) | 57 (62.6)  34 (37.4) | <0.001* |
| **Recurrence, N (%)**  No  Yes | 104 (90.4)  11 (9.6) | 50 (62.5)  30 (37.5) | <0.001* | 108 (90.8)  11 (9.2) | 46 (60.5)  30 (39.5) | <0.001* | 97 (93.3)  7 (6.7) | 57 (62.6)  34 (37.4) | <0.001* |
| *represents a statistically significant result, where p-value < 0.05; TNM, tumor, node, metastases; VATS, video-assisted thoracoscopic surgery; SCC, squamous Cell Carcinoma; AC, adenocarcinoma; LN, lymph node; CART: Classification and Regression Tree. | | | | | | | | | |

| **Supplementary Table 2.** Comparison of patient characteristics between high-risk and low-risk recurrence groups in the test cohort, defined using CART-derived cutpoints | | | | | | | | | |
| --- | --- | --- | --- | --- | --- | --- | --- | --- | --- |
| **Patient Characteristics** | **Habitat Radiomics Model** | | | **Intratumoral Radiomics Model** | | | **Combined Model** | | |
|  | **Low-risk**  **N = 64** | **High-risk**  **N = 34** | **p-value** | **Low-risk**  **N = 66** | **High-risk**  **N = 32** | **p-value** | **Low-risk**  **N = 55** | **High-risk**  **N = 43** | **p-value** |
| **Mean Age** ± **SD** | 67.20 (10.95) | 67.68 (8.67) | 0.82 | 67.18 (10.62) | 67.75 (9.34) | 0.79 | 66.64 (11.11) | 68.30 (8.87) | 0.41 |
| **Sex, N (%)**  Female  Male | 33 (51.6)  31 (48.4) | 20 (58.8)  14 (41.2) | 0.64 | 40 (60.6)  26 (39.4) | 13 (40.6)  19 (59.4) | 0.10 | 31 (56.4)  24 (43.6) | 22 (51.2)  21 (48.8) | 0.76 |
| **Race, N (%)**  White  Other | 61 (95.3)  3 (4.7) | 33 (97.1)  1 (2.9) | 0.99 | 63 (95.5)  3 (4.5) | 31 (96.9)  1 (3.1) | 0.99 | 53 (96.4)  2 (3.6) | 41 (95.3)  2 (4.7) | 0.99 |
| **Smoking History, N (%)**  Never  Current/Former | 4 (6.2)  60 (93.8) | 4 (11.8)  30 (88.2) | 0.57 | 6 (9.1)  60 (90.9) | 2 (6.2)  30 (93.8) | 0.93 | 5 (9.1)  50 (90.9) | 3 (7.0)  40 (93.0) | 0.99 |
| **TNM Stage, N (%)**  1A  1B  2A  2B  3A | 38 (59.4)  12 (18.8)  4 (6.2)  3 (4.7)  7 (10.9) | 4 (11.8)  8 (23.5)  6 (17.6)  8 (23.5)  8 (23.5) | <0.001* | 36 (54.5)  10 (15.2)  6 (9.1)  6 (9.1)  8 (12.1) | 6 (18.8)  10 (31.2)  4 (12.5)  5 (15.6)  7 (21.9) | 0.02* | 34 (61.8)  9 (16.4)  4 (7.3)  3 (5.5)  5 (9.1) | 8 (18.6)  11 (25.6)  6 (14.0)  8 (18.6)  10 (23.3) | <0.001* |
| **Surgery Type, N (%)**  Lobectomy  Segmentectomy  VATS  Other | 48 (75.0)  11 (17.2)  4 (6.2)  1 (1.6) | 25 (73.5)  2 (5.9)  1 (2.9)  6 (17.6) | 0.014* | 50 (75.8)  11 (16.7)  3 (4.5)  2 (3.0) | 23 (71.9)  2 (6.2)  2 (6.2)  5 (15.6) | 0.08 | 42 (76.4)  10 (18.2)  3 (5.5)  0 (0.0) | 31 (72.1)  3 (7.0)  2 (4.7)  7 (16.3) | 0.01* |
| **Adjuvant Therapy, N (%)**  None  Chemotherapy  Radiation or Chemoradiation | 54 (84.4)  6 (9.4)  4 (6.2) | 13 (38.2)  17 (50.0)  4 (11.8) | <0.001* | 50 (75.8)  12 (18.2)  4 (6.1) | 17 (53.1)  11 (34.4)  4 (12.5) | 0.08 | 47 (85.5)  6 (10.9)  2 (3.6) | 20 (46.5)  17 (39.5)  6 (14.0) | <0.001* |
| **Tumor Site**  Main Bronchus or Overlapping Lesion  Upper Lobe  Middle Lobe  Lower Lobe | 2 (3.1)  34 (53.1)  2 (3.1)  26 (40.6) | 2 (5.9)  18 (52.9)  1 (2.9)  13 (38.2) | 0.93 | 3 (4.5)  36 (54.5)  3 (4.5)  24 (36.4) | 1 (3.1)  16 (50.0)  0 (0.0)  15 (46.9) | 0.53 | 1 (1.8)  29 (52.7)  2 (3.6)  23 (41.8) | 3 (7.0)  23 (53.5)  1 (2.3)  16 (37.2) | 0.61 |
| **Histology, N (%)**  SCC  AC  Other | 19 (29.7)  43 (67.2)  2 (3.1) | 9 (26.5)  22 (64.7)  3 (8.8) | 0.47 | 13 (19.7)  50 (75.8)  3 (4.5) | 15 (46.9)  15 (46.9)  2 (6.2) | 0.02 | 13 (23.6)  40 (72.7)  2 (3.6) | 15 (34.9)  25 (58.1)  3 (7.0) | 0.31 |
| **Regional LN Metastases, N (%)**  No  Yes | 53 (82.8)  11 (17.2) | 17 (50.0)  17 (50.0) | 0.001* | 48 (72.7)  18 (27.3) | 22 (68.8)  10 (31.2) | 0.86 | 45 (81.8)  10 (18.2) | 25 (58.1)  18 (41.9) | 0.02* |
| **Recurrence, N (%)**  No  Yes | 58 (90.6)  6 (9.4) | 20 (58.8)  14 (41.2) | <0.001* | 58 (87.9)  8 (12.1) | 20 (62.5)  12 (37.5) | 0.01* | 52 (94.5)  3 (5.5) | 26 (60.5)  17 (39.5) | <0.001* |
| *represents a statistically significant result, where p-value < 0.05; TNM, tumor, node, metastases; VATS, video-assisted thoracoscopic surgery; SCC, squamous Cell Carcinoma; AC, adenocarcinoma; LN, lymph node; CART: Classification and Regression Tree. | | | | | | | | | |

**Supplemental Figures**

**Figure S1**

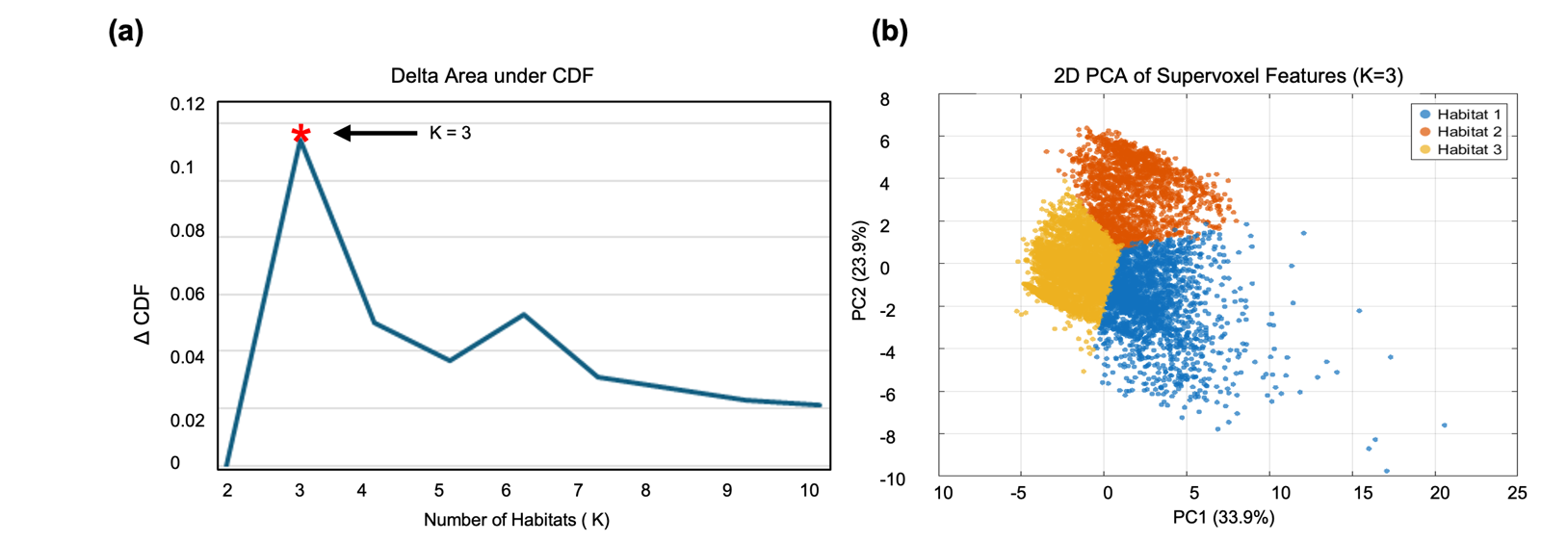

**Figure S1.** Population-level habitat number selection and visualization of supervoxel feature clustering. (a) Delta area under the consensus cumulative distribution function (CDF) plotted across different numbers of habitats (K = 2–10), demonstrating the largest incremental gain at K = 3, which was selected as the optimal number of habitats.(b) Three-dimensional principal component analysis (PCA) of supervoxel-level radiomic features for K = 3, illustrating the separation of the three identified habitats in feature space.

**Figure S2**

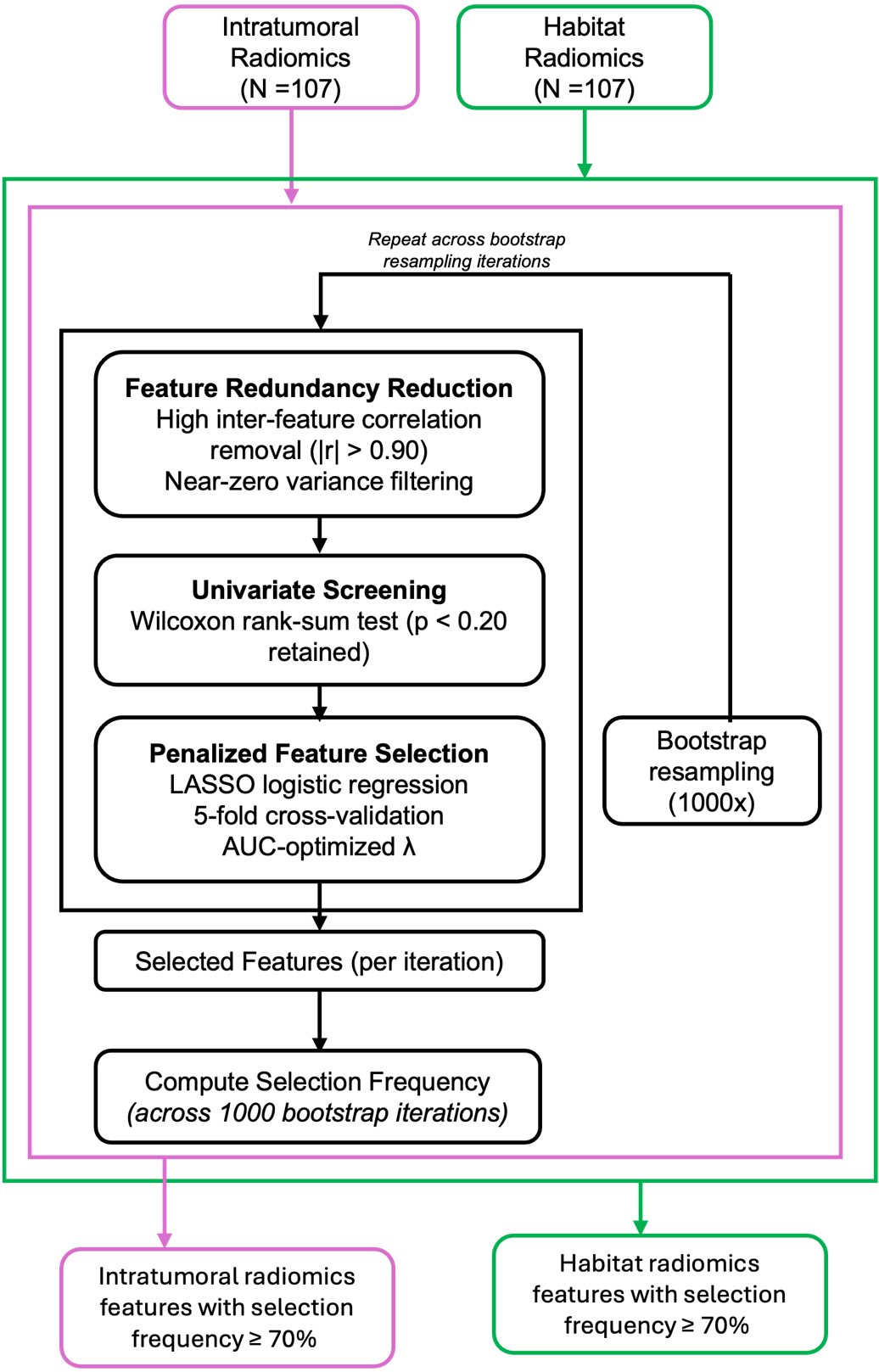

**Figure S2.** Feature selection workflow for intratumoral and habitat radiomics models. Radiomic features extracted from intratumoral and habitat-based analyses (N = 107 each) were processed using an identical, multistep feature selection pipeline. Feature redundancy was first reduced by removing highly correlated features (|r| > 0.90) and near-zero variance features. Univariate screening was then performed using the Wilcoxon rank-sum test, retaining features with p < 0.20. Penalized feature selection was subsequently conducted using LASSO logistic regression with 5-fold cross-validation and an AUC-optimized regularization parameter (λ). This process was repeated across 1000 bootstrap resampling iterations to estimate feature selection frequency. Features selected in ≥70% of bootstrap iterations were retained for downstream modeling.
